# Long term follow-up of participants in the Taking Charge after Stroke randomised controlled trial

**DOI:** 10.1101/2025.06.10.25329381

**Authors:** Alexander Martin, Vivian Fu, Zamir Joya, Sajida Joya, Allie Eathorne, Mark Weatherall, Gabrielle Shortt, Alex Semprini, John Gommans, Harry McNaughton

**Author notes:** Corresponding Author: Harry McNaughton, Medical Research Institute of New Zealand, Wellington, New Zealand. Trial Registration: UTN: U1111-1273-6693, ANZCTRN: ACTRN12622000311752.

## Abstract

**Background:** The Take Charge intervention – a conversation-based, community intervention to improve motivation, improved independence and physical health 12 months after stroke in two randomised controlled trials with 572 participants. This paper reports long term outcomes for the 400 participants in the Taking Charge After Stroke (TaCAS) study.

**Method:** A follow-up study, of a multicentre, randomised, controlled, parallel-group trial. Outcome measures were collected by postal questionnaire or by telephone call. The TaCAS study recruited 400 participants discharged after stroke, randomised within 16 weeks to one of three groups: one session of the Take Charge intervention, two sessions six weeks apart or no sessions (control). This study is of participants still alive and willing to answer a questionnaire between 5 and 6 years after their index stroke. The primary outcome was the Physical Component Summary (PCS) of the Short Form 36. Secondary outcomes were: Frenchay Activities Index; modified Rankin Score (mRS); survival; and stroke recurrence.

**Results:** Mortality data were available for all 400 participants and functional data for 204/296 (69%) of survivors. The mean difference (95% CI) in PCS between Take Charge and control groups was 2.8 (−0.8 to 6.5) units, p = 0.12, and for independence (mRS 0-2) the odds ratio (95% CI) was 0.56 (0.28 to 1.16) p = 0.11, both favouring Take Charge with similar point estimates to those after 12 months. Point estimates for other outcomes also favoured Take Charge but were not statistically significant.

**Conclusions:** Differences in physical health and independence observed at 12 months, were sustained 5-6 years after stroke, but were not statistically significant.

## Introduction

The Take Charge intervention, a conversation-based intervention designed to improve intrinsic motivation, delivered in the early community phase of stroke rehabilitation, improves independence, advanced activities of daily living and physical health at 12 months. Two randomised controlled trials of size 172 and 400 participants, and an individual participant meta-analysis of all 572 participants, provide strong evidence of benefit with a large effect size. The number needed to treat (NNT) to have one extra person independent at 12 months after stroke is eight.[1, 2] This is of a similar magnitude to that for intravenous thrombolysis with alteplase in the first 3 hours after stroke.[3] Take Charge is the only intervention that has been shown to affect these outcomes in people discharged from hospital after stroke. It is also very cost-effective, saving $US2,000 for every patient treated when assessed directly over a 12-month horizon following stroke.[4] The Take Charge intervention is now a strong recommendation in recent stroke rehabilitation guidelines [5] and, to our knowledge, is being used in clinical services in the U.S., Canada, Australia, New Zealand, U.K., Germany and other countries. Materials are free to download and available in seven languages.[6]

Long term stroke follow-up studies (>2 years after randomisation) of randomised controlled trials (RCT) are rare. This reflects the difficulties in maintaining contact with participants over long periods. Nevertheless, these follow-up data are important, to assess whether established medium-term benefits of an intervention are sustained in the longer term. To our knowledge only one other study has assessed functional outcomes in participants who received a randomised intervention multiple years after the initial trial. That study report five and 10-year survival and functional outcomes for survivors of the original 220 participants in a randomised trial of stroke unit versus general medical ward care. In that study, beneficial survival and functional outcome observed after 12 months were sustained at both five and 10 years.[7-9].

This study aimed to assess independence, physical health, and advanced activities of daily living, between five and six years after the initial stroke to explore if the positive outcomes for Take Charge participants after 12 months in the Taking Charge after Stroke (TaCAS) study were sustained.

## Methods

This is a cross-sectional follow-up study of the 400 participants enrolled in the TaCAS Study, a multicentre randomised controlled parallel-group trial in people discharged to a community setting following hospital admission with acute stroke.[2, 10]. The study protocol was approved by the Health and Disability Ethics Committee. Participants in the initial study were not masked as to treatment allocation. Outcomes for this study were collected remotely, by investigators masked to the original treatment allocation. Questionnaires were administered either by pen and paper through the post, or by telephone with an investigator.

### Participants

In the TaCAS study, 400 non-Māori, non-Pacific adults (>16 years) discharged to a community setting after acute stroke, and randomised between two and 16 weeks after that event, were recruited over 2016/2017 from seven centers across New Zealand, four tertiary and three non-tertiary centres. Of the initial 400 participants recruited, 2 withdrew and 10 died before the final visit at 12 months, with a 12-month follow-up rate for functional outcomes of 97%. Participants in this follow-up study were survivors who could be contacted and consented to complete a questionnaire.

### Sample Size

This was a convenience sample, determined by the original study size and the proportion of survivors willing to participate.

### Interventions

No new intervention occurred as part of this follow-up study. The original interventions are described in detail in the TaCAS study main report.[2] In the initial study participants were randomly allocated to one of three treatment groups, one Take Charge Session (TCS), two TCS, 6 weeks apart and no TCS in a 1:1:1 ratio. All participants received usual community rehabilitation.

### Outcome variables

The primary outcome was the Physical Component Summary (PCS) of the Short Form 36 (SF-36) adapted to New Zealand norms.[11] Secondary outcomes were the Frenchay Activities Index, the modified Rankin Score dichotomised to independent (mRS 0-2) and dependent (mRS >2), survival and stroke recurrence. All outcome measures were used in the TaCAS study (see Supplementary Table 1 for list of measures and timepoints for the original and current study). All outcome measures have been validated for use in stroke. [12-14]

### Data collection

Data were collected from March to August of 2022, between 5 and 6 years after the initial stroke. Death was established by accessing medical records. Any participants who were not listed as deceased were sent a consent form and questionnaire which they could complete and return by post. Alternatively, participants could choose a telephone call from an investigator during which informed consent and study questionnaires were completed. If a participant was unable to provide the necessary information, a carer or family member could complete the mRS alone, either in writing or by telephone on behalf of the participant. If there was no response from a participant, they were contacted by telephone between 14 and 21 days after their study pack was posted. If uncontactable, participants were subsequently contacted a second and final time between 14 and 21 days after the first follow-up. Failing this contact the participant was classified as lost to follow-up.

### Data management and missing data

Source data from paper questionnaires were transcribed by an investigator into a secure REDCap [15] database and electronic copies of the source data were uploaded alongside. Verbally collected data were input directly into this database. The REDCap database was hosted on Medical Research Institute of New Zealand (MRINZ) servers.

### Statistical analysis

Statistical analyses were prespecified in the study protocol and were intended to match the analyses of the initial study. All analyses were intention to treat. The primary outcome measure was the Physical Component Summary (PCS) of the SF-36. The primary analysis of the primary outcome was ANOVA; all Take Charge vs control. The number of Take Charge sessions was used in a sensitivity analysis to explore a dose-response relationship with the outcome. The secondary analysis of the PCS of the SF-36 was ANCOVA, adjusted for the following baseline variables: Barthel Index score three days post stroke; PCS of the Short Form 12; age; gender; living alone. The two pre-specified comparisons were: all TCS vs control; and two sessions TCS vs one session TCS. The analysis of the FAI was ANOVA. mRS, survival and stroke recurrence were analysed by logistic regression. SAS version 9.4 was used.

## Results

The mean (SD) time between initial stroke and consent for this follow-up was 5.7 (0.4) years. Mortality data were available for all the original 400 participants of the TaCAS trial. Of these, two withdrew, and 10 died by the 12-month follow-up. Of the 388 participants who completed the final visit of the original study, a further 91 died before the 5 to 6 years follow-up (25% mortality overall). Of the remaining 297 participants, 47 chose not to participate and 46 were lost to follow-up, leaving 204 (69%) of the surviving participants, who provided functional outcome data, with 191 (64%) able to complete the PCS. Loss to follow-up was similar across the three arms of the trial. The participant flow is shown in Figure 1. Baseline characteristics of participants at the 5-to-6-year follow-up were broadly similar across the intervention groups, shown in Table 1.

**Table 1:** Baseline characteristics of follow-up study participants.

|  | **Treatment group Mean (SD)** |  |  |
| --- | --- | --- | --- |
|  | **All** | **Control** | **All Take Charge** |
| Age (years) at randomisation | 69.3 (11.2) | 69.9 (11.7) | 69.1 (10.9) |
| Time (years) from stroke | 5.7 (0.4) | 5.7 (0.5) | 5.7 (0.4) |
| BI at randomisation | 15.7 (5.4) | 15.3 (5.4) | 15.9 (5.4) |
| PCS SF12 at baseline | 41.8 (7.8) | 41.1 (9.0) | 42.1 (7.2) |
|  | N (%) |  |  |
| Living alone prior to stroke | 68 (33) | 25 (42) | 43 (29) |

BI = Barthel Index; PCS = Physical Component Summary; SF12 = Short Form 12

**Figure 1:**
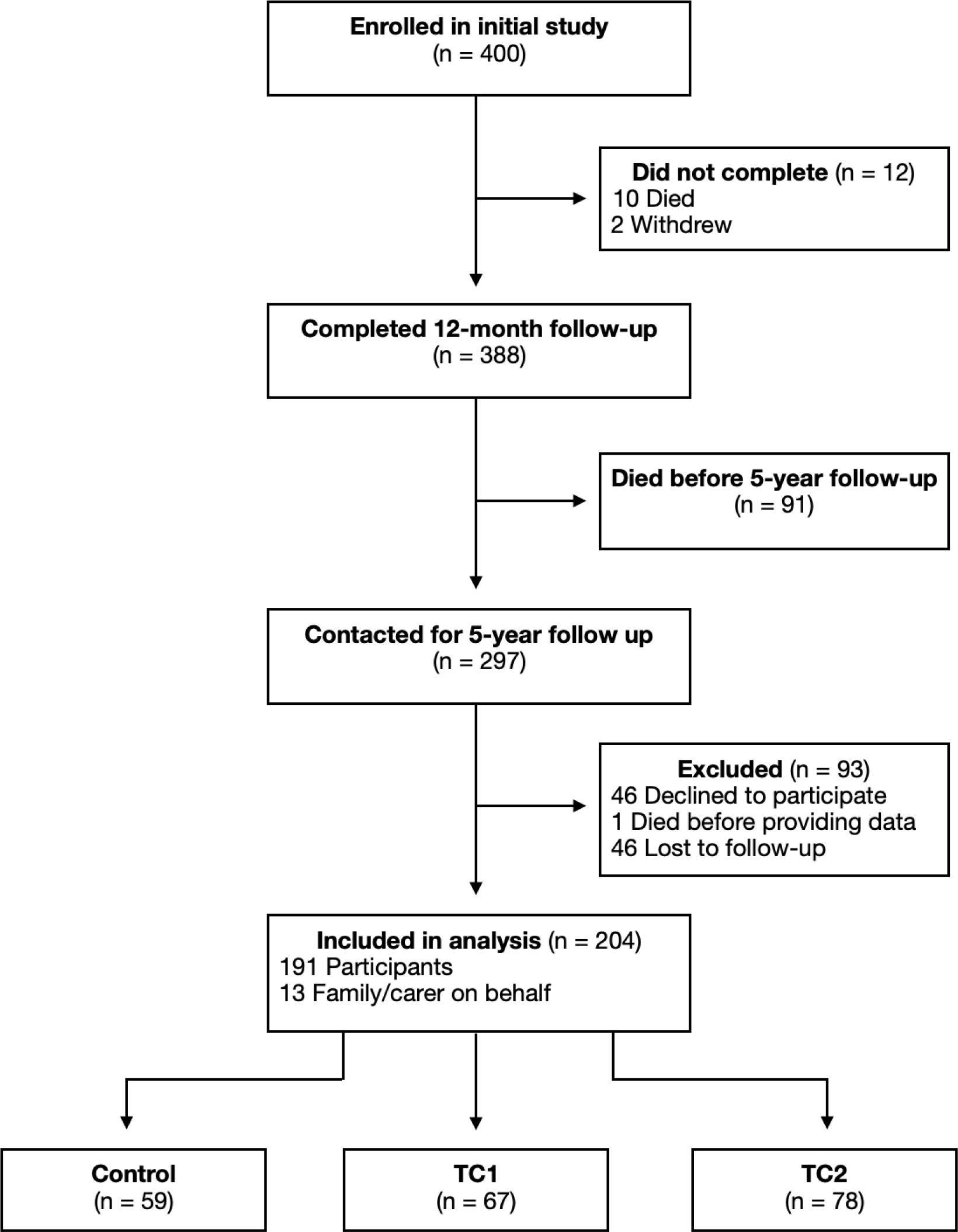
Flow of participants in Taking Charge after Stroke (TaCAS) trial, completed after 12 month follow-up, and Taking Charge after Stroke Follow up (TaCAS-FU) trial. TC1 = one Take Charge session only, TC2 = 2 Take Charge sessions

**Figure 2:**
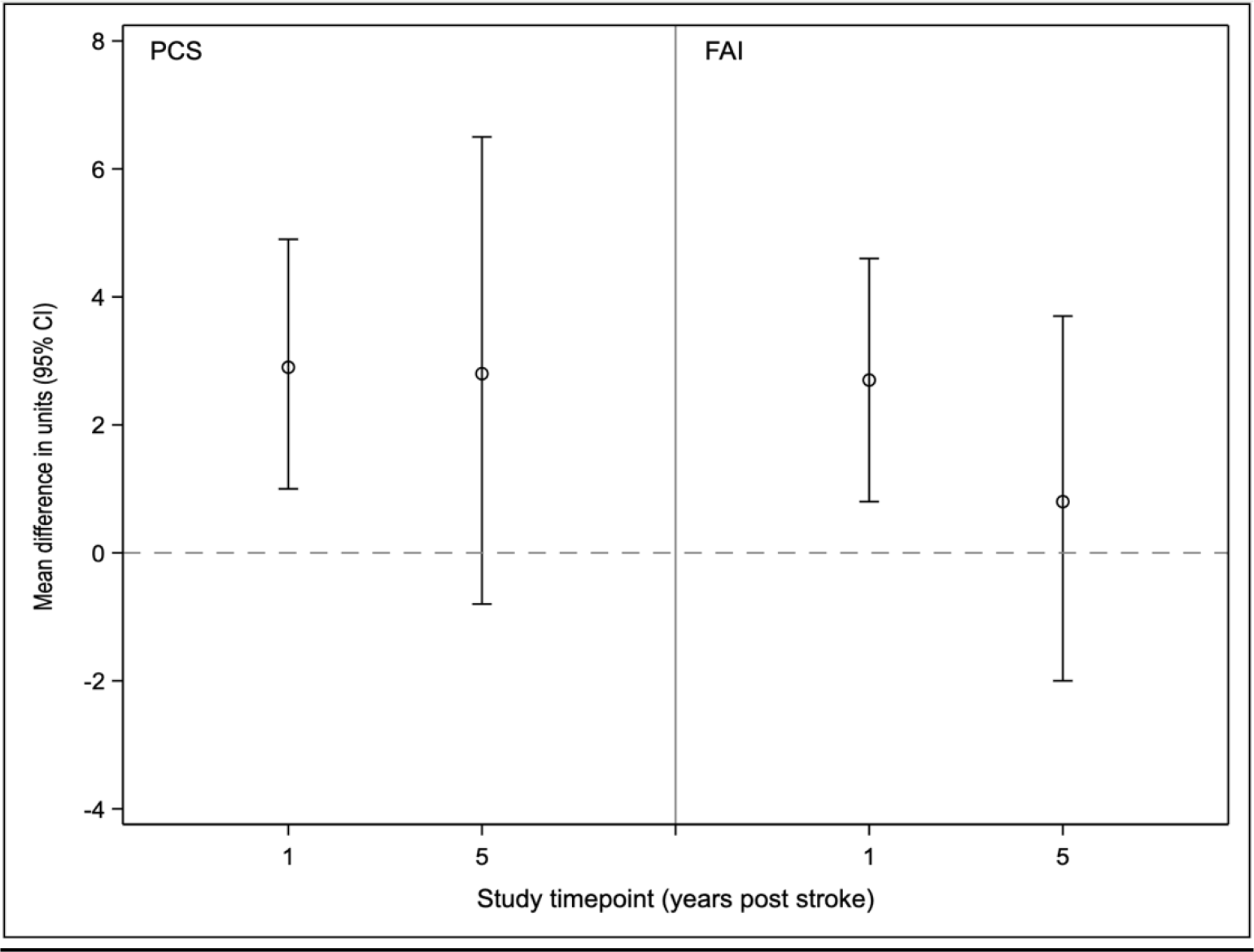
Difference between Take Charge and control group in PCS and FAI score at 12 months and 5-6 years post stroke, shown using point estimates of the mean difference in units (for PCS and FAI) and 95% confidence intervals.

**Figure 3:**
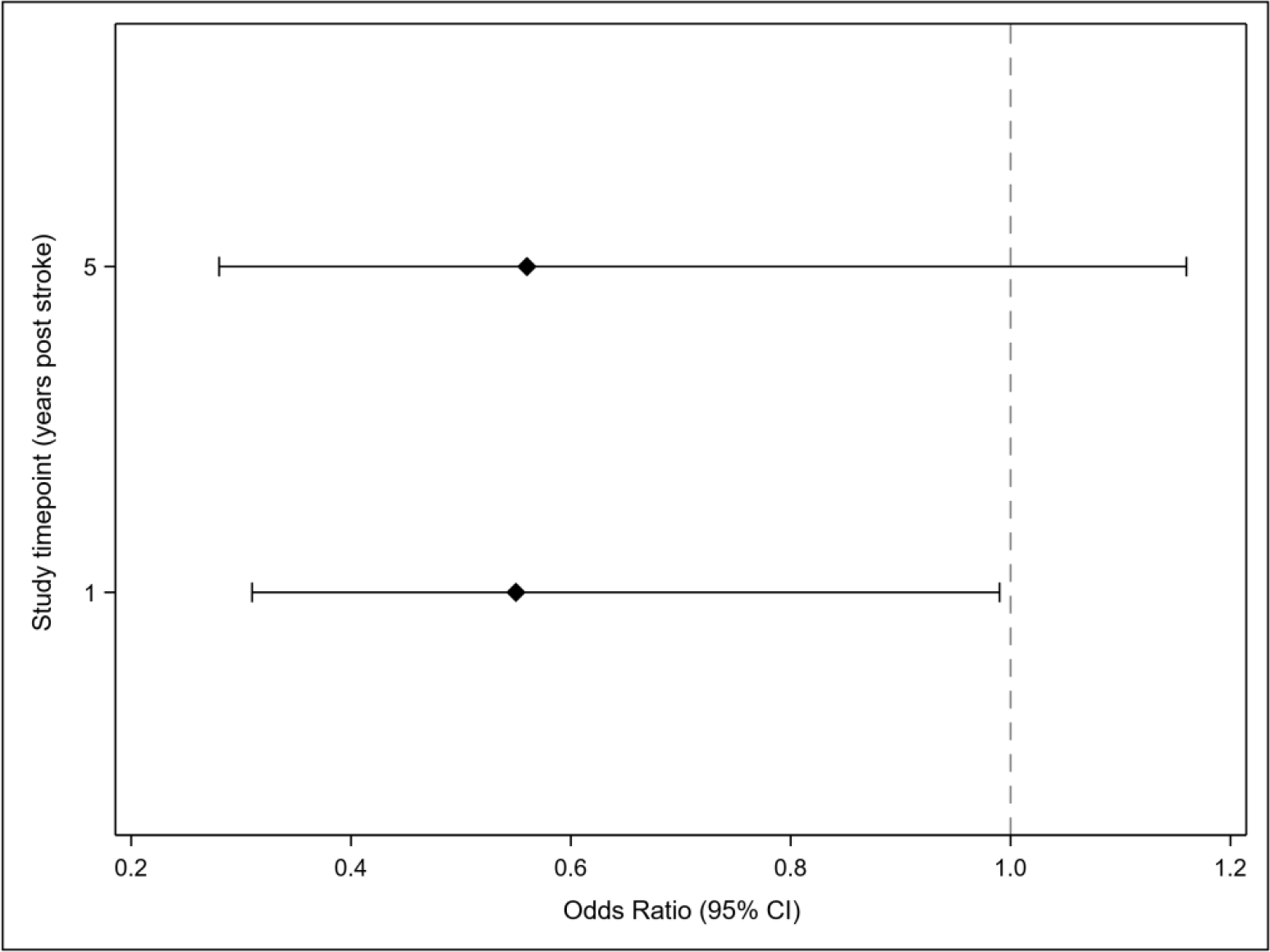
Odds ratios and 95% confidence intervals of being dependent (modified Rankin Score >2), Take Charge vs control 12 months, and between 5 and 6 years post stroke. An odds ratio below 1.0 favours Take Charge.

For the entire cohort, physical health declined from a mean PCS of 45.4 units at 12 months to 40.3 units after between 5 and 6 years of follow-up. Similarly, rates of independence (mRS 0-2) fell from 86% at 12 months to 78% after between 5 and 6 years. There were 21/189 (11%) respondents reporting a recurrent stroke.

Outcomes by treatment allocation are summarised in Table 2. Participants who received any Take Charge sessions had mean (95% CI) PCS score 2.8 units (−0.8 to 6.5), p = 0.12, higher (better) at prolonged follow-up after stroke than participants who received no Take Charge sessions. This point estimate of the mean difference after between 5 and 6 years is almost identical to that observed at 12-months after stroke, although no longer statistically significant. Similarly, the odds ratio (95% CI) for dependency (mRS >2) for Take Charge participants compared to control was 0.56 (0.28 to 1.16), p=0.11, at the prolonged follow-up, as compared to an odds ratio of 0.55 (0.31 to 0.99), p = 0.045, on the same measure 12 months after stroke. An odds ratio less than one favours the Take Charge intervention. The mean difference (95% CI) between groups in the FAI was 0.8 (−2.0 to 3.7) p = 0.57, favouring Take Charge. Fewer participants in the Take Charge groups than control were dead or reported recurrent stroke by 5 to 6 years, however the differences were small and not statistically significant. Mean (95% CI) PCS scores increased 1.6 units (−0.4 to 3.6), p = 0.11, for each extra TC session received. This is a similar estimate of change to that measured at 12 months, which showed an increase of 1.9 (0.8 to 3.1), p<0.001, units per extra session received.

**Table 2:** Outcomes between 5 and 6 years compared to outcomes 12 months after stroke.

|  | 5-6 Years After Stroke |  |  |  | 12-Months After Stroke |  |  |  |
| --- | --- | --- | --- | --- | --- | --- | --- | --- |
|  | All N=191 | Control<br>N=55 | All TC<br>N=136 | TC-Control | All<br>N=386 | Control<br>N=130 | All TC<br>N=256 | TC-Control |
|  | Mean (SD) |  |  | Estimate<br>(95% CI) | Mean (SD) |  |  | Estimate<br>(95% CI) |
| PCS | 40.3<br>(11.6) | 38.3 (13) | 41.2<br>(10.9) | 2.8<br>(-0.8 to 6.5)<br>p = 0.12 | 45.4<br>(8.2) | 43.4<br>(10.7) | 46.4 (8.4) | 2.9<br>(0.95 to 4.9)<br>p = 0.004 |
| FAI | 30.6<br>(9.1) | 30 (10.4) | 30.8<br>(8.6) | 0.8<br>(-2.0 to 3.7)<br>p = 0.57 | 30.6<br>(9.1) | 26.0<br>(10.0) | 28.7 (8.5) | 2.7<br>(0.8 to 4.6)<br>p = 0.006 |
|  | N/N (%) |  |  | Odds ratio<br>(95% CI) | N/N (%) |  |  | Odds ratio<br>(95% CI) |
| Alive | 297/400<br>(74) | 94/130<br>(72) | 203/270<br>(75) | 1.31<br>(0.80 to 2.12)<br>p = 0.28 | 390/400<br>(98) | 128/130<br>(99) | 262/270<br>(97) | 1.95<br>(0.41 to 9.34)<br>p = 0.40 |
| mRS<br><2 <sup>a</sup> | 162/203<br>(80) | 43/59<br>(73) | 119/144<br>(83) | 0.56<br>(0.28 to 1.16)<br>p = 0.11 | 331/387<br>(86) | 103/128<br>(81) | 228/259<br>(88) | 0.55<br>(0.31 to 0.99)<br>p = 0.045 |
| Recur<br>rent<br>stroke | 21/189<br>(11) | 7/55 (13) | 14/134<br>(10) | 0.80<br>(0.30 to 2.1)<br>p = 0.64 | 24/395<br>(6) | 10/130 (8) | 14/265 (5) | 0.61<br>(0.25 to 1.45)<br>p = 0.26 |

a: the odds ratio is for dependence (mRS >2), meaning a number less than one favours Take Charge
TC = Take Charge; PCS = Physical Component Summary; FAI = Frenchay Activities Index; mRS = Modified Rankin Score

## Discussion

In this long term follow-up of a randomised trial after stroke, we were able to assess mortality for all the original participants, and data regarding functional outcomes for 69% of survivors, a mean 5.7 years after the original stroke. The main finding was that the differences in physical health outcomes and independence, favouring the Take Charge intervention, measured at 12 months after stroke, were sustained at almost 6 years of follow-up. However, these differences were no longer statistically significant, reflecting participant attrition from mortality and loss to follow-up. The measured 2.8 units difference in physical health, approximates the 2.5-3 unit minimal clinically important difference (MCID) for the PCS in people with stroke.[16] A dose-response effect was again observed, with higher PCS scores for participants receiving two sessions compared to one.

To our knowledge this is the largest long-term follow-up study of a randomised intervention for stroke. Our results show a similar pattern to the stroke unit studies of Indredavik et al (7-9) where a difference observed at 12 months persisted to 5 (and 10) years. Those studies were numerically smaller than ours with functional outcomes limited to a simple activity of daily living measure, the Barthel Index (treated as a dichotomous variable), obtained from 58 participants at 5 years and 41 participants at 10 years. Our functional data were based on responses from 204 individuals and measured physical health, advanced activities of daily living and independence giving a broader picture of outcomes at International Classification of Functioning levels more appropriate for people living in the community.[17] The confirmation that significant changes at 12 months could be sustained long-term is important for at least two reasons. Firstly, it should encourage attempts to maximise outcomes in the first year after stroke and secondly, the information can be used in longer term modelling of outcomes and cost effectiveness of interventions after stroke.

The most significant limitation of the study was loss to follow-up which resulted in a smaller achieved sample size and possible Type II error. Nevertheless, we were able to obtain complete survival data and functional data were obtained from over two-thirds of survivors. Participants in the initial study were not masked to treatment allocation and participants with more favourable outcomes may have been more likely to respond to the request for follow-up. However, loss to follow-up was distributed evenly between groups and the similarity in the differences at 12 months and 5-6 years between the groups is compelling.

That a low intensity (one or two sessions) conversation-based intervention delivered in the first four months after stroke could have a significant effect on independence and physical health 12 months after stroke may be surprising but is supported by sound theory and experimental evidence.[18]. That the effect could be sustained past 5 years of follow-up seems remarkable. We hypothesise that the Take Charge intervention influences intrinsic motivation of the individual leading to permanent behaviour change, with the person with stroke taking more responsibility for achieving the outcomes that matter most to them.[18-20]

These results strengthen the case for widespread implementation of the Take Charge intervention for people discharged home after stroke. They also underline the potential for psychological interventions in general after stroke to have an important impact on long term recovery, when combined with usual therapy-based approaches.[18]

## Data Availability

Data are available on reasonable request from the corresponding author

## Acknowledgements

Thanks to the participants in the original Taking Charge after Stroke study and the large number who also agreed to participate in this follow up study.

